# Outcomes of laboratory-confirmed SARS-CoV-2 infection during resurgence driven by Omicron lineages BA.4 and BA.5 compared with previous waves in the Western Cape Province, South Africa

**DOI:** 10.1101/2022.06.28.22276983

**Authors:** Mary-Ann Davies, Erna Morden, Petro Rosseau, Juanita Arendse, Jamy-Lee Bam, Linda Boloko, Keith Cloete, Cheryl Cohen, Nicole Chetty, Pierre Dane, Alexa Heekes, Nei-Yuan Hsiao, Mehreen Hunter, Hannah Hussey, Theuns Jacobs, Waasila Jassat, Saadiq Kariem, Reshma Kassanjee, Inneke Laenen, Sue Le Roux, Richard Lessells, Hassan Mahomed, Deborah Maughan, Graeme Meintjes, Marc Mendelson, Ayanda Mnguni, Melvin Moodley, Katy Murie, Jonathan Naude, Ntobeko A. B. Ntusi, Masudah Paleker, Arifa Parker, David Pienaar, Wolfgang Preiser, Hans Prozesky, Peter Raubenheimer, Liezel Rossouw, Neshaad Schrueder, Barry Smith, Mariette Smith, Wesley Solomon, Greg Symons, Jantjie Taljaard, Sean Wasserman, Robert J. Wilkinson, Milani Wolmarans, Nicole Wolter, Andrew Boulle, the Western Cape Department of Health and Wellness and the South African National Departments of Health in collaboration with the National Institute for Communicable Diseases in South Africa

**Affiliations:** Health Intelligence, Western Cape Government: Health and Wellness, South Africa; Centre for Infectious Disease Epidemiology and Research, School of Public Health and Family Medicine, University of Cape Town, South Africa; Division of Public Health Medicine, School of Public Health and Family Medicine, University of Cape Town, South Africa; National Department of Health, South Africa; Western Cape Government: Health and Wellness, South Africa; Groote Schuur Hospital, Western Cape Government: Health and Wellness, South Africa; Division of Infectious Diseases and HIV Medicine, Department of Medicine, University of Cape Town, South Africa; National Institute for Communicable Diseases, National Health Laboratory Service, South Africa; School of Public Health, Faculty of Health Sciences, University of the Witwatersrand, Johannesburg, South Africa; Division of Medical Virology, University of Cape Town, Cape Town, Western Cape, South Africa; National Health Laboratory Service, South Africa; Metro Health Services, Western Cape Government: Health and Wellness; Division of Health Systems and Public Health, Department of Global Health, Faculty of Medicine and Health Sciences, Stellenbosch University; Karl Bremer Hospital, Western Cape Government: Health and Wellness; KwaZulu-Natal Research, Innovation & Sequencing Platform, University of KwaZulu-Natal, Durban, South Africa; Department of Medicine, University of Cape Town, South Africa; Khayelitsha District Hospital, Western Cape Government: Health and Wellness; Mitchells Plain Hospital, Western Cape Government: Health and Wellness; South African Medical Research Council Extramural Unit on Intersection of Noncommunicable Diseases and Infectious Diseases; Tygerberg Hospital, Western Cape Government: Health and Wellness; Division of Infectious Diseases, Department of Medicine, Stellenbosch University, South Africa; Rural Health Services, Western Cape Government: Health and Wellness; Division of Medical Virology, University of Stellenbosch, South Africa; Division of General Medicine, Department of Medicine, Stellenbosch University, South Africa; The Francis Crick Institute, Midland Road, London, NW1 1AT, UK; Department of Infectious Diseases, Imperial College London, W12 0NN, UK; Wellcome Centre for Infectious Disease Research in Africa, Institute of Infectious Disease and Molecular Medicine, University of Cape Town, South Africa; School of Pathology, Faculty of Health Sciences, University of Witwatersrand, Johannesburg, South Africa

**Keywords:** COVID-19, Omicron, BA.1, BA.4, BA.5, vaccination, prior infection, death, severe hospitalization

## Abstract

**Objective:** We aimed to compare clinical severity of Omicron BA.4/BA.5 infection with BA.1 and earlier variant infections among laboratory-confirmed SARS-CoV-2 cases in the Western Cape, South Africa, using timing of infection to infer the lineage/variant causing infection.

**Methods:** We included public sector patients aged ≥20 years with laboratory-confirmed COVID-19 between 1-21 May 2022 (BA.4/BA.5 wave) and equivalent prior wave periods. We compared the risk between waves of (i) death and (ii) severe hospitalization/death (all within 21 days of diagnosis) using Cox regression adjusted for demographics, comorbidities, admission pressure, vaccination and prior infection.

**Results:** Among 3,793 patients from the BA.4/BA.5 wave and 190,836 patients from previous waves the risk of severe hospitalization/death was similar in the BA.4/BA.5 and BA.1 waves (adjusted hazard ratio [aHR] 1.12; 95% confidence interval [CI] 0.93; 1.34). Both Omicron waves had lower risk of severe outcomes than previous waves. Prior infection (aHR 0.29, 95% CI 0.24; 0.36) and vaccination (aHR 0.17; 95% CI 0.07; 0.40 for boosted vs. no vaccine) were protective.

**Conclusion:** Disease severity was similar amongst diagnosed COVID-19 cases in the BA.4/BA.5 and BA.1 periods in the context of growing immunity against SARS-CoV-2 due to prior infection and vaccination, both of which were strongly protective.

## Background

The Omicron SARS-CoV-2 variant of concern (VOC) has been dominant globally since November 2021, with several lineages causing surges in infections (Iketani et al., 2022, Tegally et al., 2022, Viana et al., 2022). South Africa experienced an initial large BA.1 infection surge from November 2021 to January 2022. BA.1 was then replaced by BA.2 but with no increase in cases numbers, and this was followed by a BA.4/BA.5 infection surge between April and June 2022 (Tegally et al., 2022, Viana et al., 2022). The combination of mutations in BA.4/BA.5 appear to confer a growth advantage over BA.2, as well as immune escape from vaccine-derived and BA.1 elicited antibodies (Khan et al., 2022, Tegally et al., 2022). A growing number of BA.4 and BA.5 infections are now being reported globally (Callaway, 2022, UK Health Security Agency, 2022).

We therefore compared outcomes of laboratory-confirmed SARS-CoV-2 infections during the recent resurgence (proxy for BA.4/ BA.5 infection) with outcomes during each of the four previous waves in South Africa, each of which were caused by a different variant or lineage, using data on patients with laboratory-confirmed SARS-CoV-2 infection aged ≥20 years using public sector services in the Western Cape Province, South Africa.

## Methods

We conducted a cohort study using de-identified data from the Western Cape Provincial Health Data Centre (WCPHDC) of public sector patients aged ≥20 years with a laboratory confirmed COVID-19 diagnosis (positive SARS-CoV-2 PCR or antigen test). The Western Cape has nearly 7 million inhabitants, of whom approximately 75% use public sector health services (Western Cape Department of Health, 2020). The WCPHDC and methods for this study have previously been described in detail (Boulle et al., 2019, Davies et al., 2022, Hussey et al., 2022, Western Cape Department of Health in collaboration with the National Institute for Communicable Diseases, 2020). Briefly, for this analysis, waves of infection were defined as starting and ending when the 7-day moving average of public sector COVID-19 hospital admissions exceeded and dropped below 5 and 12 per million population respectively. We included cases diagnosed from seven days before the wave start to seven days before the wave end date to account for the lag between infection/first symptoms and hospitalization. We thus included data on cases diagnosed from 1-21 May 2022 for the BA.4/BA.5 wave, with follow-up through to 11 June 2022, corresponding to the period when BA.4/BA.5 dominated in the province (Network for Genomic Surveillance in South Africa, 2022).

We used Cox regression adjusted for age, sex, geographic location, comorbidities, service pressure (number of weekly admissions in the health district) at time of diagnosis, prior diagnosed infection (≥1 laboratory confirmed SARS-CoV-2 diagnosis ≥90 days previously) and SARS-CoV-2 vaccination to assess differences in the following COVID-19 outcomes between waves driven by different variants: (i) death and (ii) death or severe hospitalization (admission to intensive care or mechanical ventilation or oral/intravenous steroid prescription). We only included outcomes within 21 days of COVID-19 diagnosis for comparable ascertainment across all waves. All deaths within 21 days of a COVID-19 diagnosis were included unless a clear non-COVID-19 cause of death was recorded. For patients with recorded South African national identity numbers, data are linked to the South African vital registry to identify deaths not recorded in the WCPHDC. Vaccination data was obtained by linking the South African national identifier to the Electronic Vaccine Data System which records all vaccines administered in the country. The only vaccines available in South Africa to date are BNT162b and Ad26.COV2.S. Vaccination status was categorized as either (i) “boosted” (three or more homologous or heterologous doses of any vaccine), (ii) “two doses” (two doses of any vaccine) or (iii) “single dose” (single dose of Ad26.COV2.S), as the latter is considered complete primary vaccination in South Africa.

The study was approved by the University of Cape Town and Stellenbosch University Health Research Ethics Committees and Western Cape Government: Health. Individual informed consent requirement was waived for this secondary analysis of de-identified data.

## Results

We included 3,793 patients diagnosed in the BA.4/BA.5 wave and 27,614 (BA.1), 68,715 (Delta), 54,268 (Beta) and 40,204 (ancestral) in waves driven by previous variants (Table 1). The proportion with prior diagnosed infection was substantially higher in the BA.4/BA.5 (18.9%) and BA.1 (11.9%) waves compared to previous waves (<3%). In the BA.4/BA.5 wave, 12.9% of COVID-19 cases had received “single dose” Ad26.COV2.S vaccination, 36.1% had received “two doses” and 6.7% were “boosted” vaccinees.

**Table 1:** Characteristics and outcomes of COVID-19 cases included from each infection period in the Western Cape

|  | Ancestral wave<br>25 Apr to 22 Jul 2020<sup>a</sup><br>(n=40,204) | Beta wave<br>3 Nov 2020 to 22 Jan 2021<sup>a</sup><br>(n=54,268) | Delta wave<br>30 May to 10 Sep 2021<sup>a</sup><br>(n=68,750) | BA.1 wave<br>27 Nov 2021 to 12 Jan 2022<sup>a</sup><br>(n=27,614) | BA.4/BA.5 wave<br>1 May to 21 May 2022<sup>a</sup><br>(n=3,793) |
| --- | --- | --- | --- | --- | --- |
| **Male sex** | 13,380 (33.3%) | 19,083 (35.2%) | 25,948 (37.7%) | 9,630 (34.9%) | 1,327 (35.0%) |
| **Age** |  |  |  |  |  |
| **20-39 years** | 18,720 (46.6%) | 21,839 (40.2%) | 29,720 (43.2%) | 13,944 (50.5%) | 1,783 (47.0%) |
| **40-49 years** | 8,280 (20.6%) | 10,594 (19.5%) | 14,163 (20.6%) | 4,905 (17.8%) | 767 (20.2%) |
| **50-59 years** | 6,982 (17.4%) | 10,493 (19.3%) | 13,294 (19.3%) | 4,216 (15.3%) | 623 (16.4%) |
| **60-69 years** | 3,733 (9.3%) | 6,929 (12.8%) | 6,780 (9.9%) | 2,554 (9.3%) | 333 (8.8%) |
| **≥70 years** | 2,489 (6.2%) | 4,413 (8.1%) | 4,793 (7.0%) | 1,995 (7.2%) | 287 (7.6%) |
| **Non-communicable diseases** |  |  |  |  |  |
| **diabetes** | 8,265 (20.6%) | 11,509 (21.1%) | 11,581 (16.9%) | 3,627 (13.1%) | 406 (10.7%) |
| **hypertension** | 13,065 (32.5%) | 19,070 (35.1%) | 21,170 (30.8%) | 7,063 (25.6%) | 842 (22.2%) |
| **chronic kidney disease** | 2,013 (5.0%) | 2,778 (5.2%) | 3,018 (4.4%) | 958 (3.5%) | 124 (3.3%) |
| **chronic pulmonary disease / asthma** | 3,099 (7.7%) | 4,661 (8.6%) | 6,434 (9.4%) | 3,040 (11.0%) | 411 (10.8%) |
| **Tuberculosis** |  |  |  |  |  |
| **previous tuberculosis** | 2,777 (6.9%) | 3,450 (6.4%) | 4,850 (7.1%) | 2,229 (8.1%) | 232 (6.1%) |
| **current tuberculosis** | 513 (1.3%) | 555 (1.0%) | 803 (1.2%) | 578 (2.1%) | 76 (2.0%) |
| **HIV positive** | 6,203 (15.4%) | 5,512 (10.2%) | 5,925 (8.6%) | 3,298 (11.9%) | 307 (8.1%) |
| **Prior diagnosed SARS-CoV-2 infection** | 0 (0%) | 618 (1.1%) | 1,798 (2.6%) | 3,179 (11.5%) | 715 (18.9%) |
| **Vaccination<sup>b</sup>** |  |  |  |  |  |
| **none** | N/A | N/A | 63,644 (92.6%) | 14,471 (52.4%) | 1,535 (40.5%) |
| **single dose Ad26.COV2.S** | N/A | N/A | 2,501 (3.6%) | 4,069 (14.7%) | 488 (12.9%) |
| **single dose BNT162b2** | N/A | N/A | 2,289 (3.3%) | 1,144 (4.1%) | 147 (3.9%) |
| **2 doses Ad26.COV2.S** | N/A | N/A | 30 (0.04%) | 1,127 (4.1%) | 298 (7.9%) |
| **2 doses BNT162b2** | N/A | N/A | 286 (0.4%) | 6,763 (24.5%) | 1,067 (28.1%) |
| **2 doses Ad26.COV2.S + BNT162b2** | N/A | N/A | N/A | N/A | 5 (0.1%) |
| **≥3 doses Ad26.COV2.S** | N/A | N/A | N/A | 36 (0.1%) | 38 (1.0%) |
| **≥3 doses BNT162b2** | N/A | N/A | N/A | 4 (0.01%) | 192 (5.1%) |
| **≥3 doses Ad26.COV2.S + BNT162b2** | N/A | N/A | N/A | N/A | 23 (0.6%) |
| **Outcomes within 21 days of diagnosis** |  |  |  |  |  |
| **severe admission (not deceased)<sup>c</sup>** | N/A<sup>c</sup> | 1,916 (3.5%) | 2,066 (3.0%) | 481 (1.7%) | 61 (1.6%) |
| **death** | 2,147 (5.3%) | 3,717 (6.9%) | 4368 (6.4%) | 699 (2.5%) | 70 (1.9%) |

<sup>a</sup>Date of diagnoses for cases included in each wave. We included cases diagnosed from 7 days prior to the "wave start" to the date of wave end (deemed to occur when 7 day moving average of daily new public sector admissions exceeded 5/million (start) and dropped below 12/million (end) respectively). <sup>b</sup>Vaccination is summarized as vaccine type and number of doses provided diagnosis was ≥28 days after first dose, ≥14 days after second dose, and ≥7 days after third dose; <sup>c</sup>Admission to an intensive care unit, mechanical ventilation or prescription of oral or intravenous steroids; not reported for wave 1 as steroids not widely used until after 16 June 2020. N/A = not applicable

The adjusted hazard of severe hospitalization or death in the BA.4/BA.5 wave was similar to the BA.1 wave (adjusted hazard ratio [aHR] 1.12; 95% confidence interval [CI]: 0.93; 1.34) (Table 2). Both Omicron-driven waves had lower hazards of severe hospitalization or death than previous waves (Table 2). Prior diagnosed infection was strongly protective against severe hospitalization or death (aHR 0.29; 95% CI 0.24; 0.36) as was vaccination with aHR (95% CI) of 0.17 (0.07; 0.40); 0.37 (0.33; 0.42) and 0.26 (0.21; 0.32) for “boosted”, “two doses” and “single dose”, respectively. In a model not adjusting for vaccination and prior diagnosed infection, the hazard of severe hospitalization or death in the BA.4/BA.5 vs. BA.1 waves was reduced compared to the fully adjusted model (aHR 0.90; 95% CI: 0.75; 1.08). In an analysis restricted to the BA.4/BA.5 period, prior diagnosed infection remained strongly protective against severe hospitalization or death (aHR 0.23; 95% CI 0.10; 0.52) as did vaccination (aHR [95% CI]: 0.20 (0.08; 0.49); 0.39 (0.25; 0.59) and 0.51 (0.27; 0.99) for “boosted”, “two doses” and “single dose”, respectively. Results were all similar when examining the outcome of death alone.

**Table 2:** Associations between different infection periods and severe COVID-19 outcomes adjusted for patient characteristics, sub-district, vaccination, and prior diagnosed infection using Cox regression.

|  | Outcome = death<br>not adjusted for<br>vaccination and prior<br>infection |  | Outcome = death<br>adjusted for vaccination<br>and prior infection |  | Outcome = severe<br>hospitalization<sup>a</sup>/death<br>not adjusted for<br>vaccination or prior<br>diagnosed infection |  | Outcome = severe<br>hospitalization<sup>a</sup>/death<br>adjusted for<br>vaccination or prior<br>diagnosed infection |  |
| --- | --- | --- | --- | --- | --- | --- | --- | --- |
|  | Adjusted<sup>b</sup><br>HR | 95% CI | Adjusted<br>HR | 95% CI | Adjusted<sup>b</sup><br>HR | 95% CI | Adjusted<br>HR | 95% CI |
| **Male sex (vs. female)** | 1.40 | 1.34; 1.45 | 1.40 | 1.34; 1.45 | 1.27 | 1.23; 1.31 | 1.26 | 1.22; 1.30 |
| **Age (vs. 20-39 years)** |  |  |  |  |  |  |  |  |
| 40-49 years | 2.54 | 2.30; 2.81 | 2.57 | 2.33; 2.84 | 2.00 | 1.87; 2.15 | 2.04 | 1.90; 2.19 |
| 50-59 years | 5.46 | 4.99; 5.97 | 5.56 | 5.08; 6.08 | 3.42 | 3.21; 3.65 | 3.50 | 3.28; 3.74 |
| 60-69 years | 12.55 | 11.47; 13.73 | 12.88 | 11.77; 14.10 | 6.39 | 5.97; 6.83 | 6.56 | 6.13; 7.01 |
| ≥70 years | 23.19 | 21.15; 25.43 | 23.93 | 21.82; 26.24 | 10.35 | 9.65; 11.09 | 10.65 | 9.94; 11.42 |
| **Comorbidities (vs. comorbidity absent)** |  |  |  |  |  |  |  |  |
| diabetes | 2.01 | 1.92; 2.10 | 2.01 | 1.93; 2.10 | 1.97 | 1.89; 2.04 | 1.98 | 1.91; 2.06 |
| hypertension | 1.08 | 1.03; 1.13 | 1.07 | 1.02; 1.12 | 1.18 | 1.14; 1.23 | 1.17 | 1.13; 1.22 |
| chronic kidney disease | 1.90 | 1.80; 2.00 | 1.90 | 1.81; 2.00 | 1.63 | 1.56; 1.70 | 1.63 | 1.56; 1.70 |
| chronic pulmonary disease / asthma | 0.98 | 0.93; 1.04 | 0.99 | 0.93; 1.04 | 1.18 | 1.13; 1.23 | 1.19 | 1.14; 1.24 |
| previous tuberculosis | 1.30 | 1.20; 1.40 | 1.28 | 1.19; 1.38 | 1.25 | 1.17; 1.33 | 1.23 | 1.16; 1.31 |
| current tuberculosis | 2.53 | 2.20; 2.91 | 2.44 | 2.13; 2.81 | 2.89 | 2.59; 3.23 | 2.79 | 2.50; 3.11 |
| HIV | 1.60 | 1.48; 1.72 | 1.60 | 1.49; 1.72 | 1.54 | 1.45; 1.64 | 1.54 | 1.45; 1.64 |
| **Number of admissions in district in week**<br>**of diagnosis (vs <1/3 of maximum)** |  |  |  |  |  |  |  |  |
| 1/3 to <2/3 | 1.11 | 1.05; 1.17 | 1.12 | 1.06; 1.18 | 1.03 | 0.98; 1.08 | 1.04 | 0.99; 1.09 |
| ≥2/3 | 1.12 | 1.05; 1.20 | 1.13 | 1.06; 1.21 | 1.05 | 0.99; 1.11 | 1.06 | 1.00; 1.12 |
| **Prior diagnosed SARS CoV-2 infection** |  |  |  |  |  |  |  |  |
| Yes (vs none) |  |  | 0.51 | 0.42; 0.63 |  |  | 0.29 | 0.24; 0.36 |
| **Vaccination (vs. None)<sup>c</sup>** |  |  |  |  |  |  |  |  |
| single dose Ad26.COV2.S |  |  | 0.24 | 0.18; 0.33 |  |  | 0.26 | 0.21; 0.32 |
| two doses (Ad26.COV2.S and/or<br>BNT162b2) |  |  | 0.36 | 0.31; 0.42 |  |  | 0.37 | 0.33; 0.42 |
| boosted (≥ 3doses Ad26.COV2.S<br>and/or BNT162b2) |  |  | 0.06 | 0.01; 0.40 |  |  | 0.17 | 0.07; 0.40 |
| **Wave period (dominant variant)** |  |  |  |  |  |  |  |  |
| wave 1 (ancestral) | 2.08 | 1.90; 2.28 | 1.30 | 1.17; 1.44 | N/A<sup>a</sup> |  | N/A<sup>a</sup> |  |
| wave 2 (Beta) | 2.35 | 2.16; 2.57 | 1.47 | 1.34; 1.62 | 2.06 | 1.93; 2.20 | 1.28 | 1.20; 1.38 |
| wave 3 (Delta) | 2.58 | 2.37; 2.81 | 1.75 | 1.59; 1.92 | 2.16 | 2.03; 2.29 | 1.44 | 1.35; 1.54 |
| wave 4 (Omicron BA.1) | Ref |  | Ref |  | Ref |  | Ref |  |
| wave 5 (Omicron BA.4/BA.5) | 0.93 | 0.72; 1.20 | 1.16 | 0.90; 1.50 | 0.90 | 0.75; 1.08 | 1.12 | 0.93; 1.34 |

<sup>a</sup>Admission to an intensive care unit, mechanical ventilation or prescription of oral or intravenous steroids; not reported for wave 1 as steroids not widely used until after 16 June 2020. <sup>b</sup>Adjusted for all variables shown in the table as well as subdistrict/district, but not for vaccination or prior diagnosed infection
<sup>c</sup>Vaccination status is categorized as “single dose” (≥28 days after single dose Ad26.COV2.S), “two doses” (≥14 days after second dose of homologous or heterologous vaccination with Ad26.COV2.S and/or BNT162b2), and “boosted” (≥7 days after third dose of homologous or heterologous vaccination with Ad26.COV2.S and/or BNT162b2); HR = hazard ratio; CI = confidence interval; N/A = not applicable

## Discussion

Using the period of diagnosis as a proxy for being infected with different Omicron lineages in the Western Cape, we found no difference in the risk of severe COVID-19 hospitalization or death during the BA.4/BA.5 period compared to the BA.1 period, both of which had better outcomes than previous waves. Strong protection against severe COVID-19 conferred by prior infection and vaccination was retained in the BA.4/BA.5 wave, with three homologous doses of Ad26.COV2.S or BNT162b2 or a heterologous combination of these providing 83% protection (95% CI 60; 93%) against severe COVID-19 hospitalization or death amongst laboratory-confirmed cases.

A study in animals recently suggested that BA.4/BA.5 may be more pathogenic than BA.2 (Kimura et al., 2022). Although we did not compare BA.4/BA.5 with BA.2 directly as BA.2 did not cause a distinguishable surge in infections in the Western Cape, disease severity of BA.2 and BA.1 are similar (Lewnard et al., 2022) and we found no evidence of worse clinical outcomes with BA.4/BA.5 compared to BA.1. Nonetheless, our findings need to be interpreted in the context of South African SARS-CoV-2 epidemiology with progressively increasing seroprevalence due to prior infection (mostly undiagnosed) and/or vaccination (Bingham et al., 2022, Madhi et al., 2022, Sun et al., 2022). For example, among blood donors, after the BA.1 wave the estimated national prevalence of anti-nucleocapsid antibodies was 87% (indicating previous infection) with a further 10% having anti-spike antibodies only (suggesting vaccination without prior infection) (Bingham et al., 2022). Indeed, our finding that the aHR shifted towards a lower risk of severe outcomes during BA.4/BA.5 vs. BA.1 in models not accounting for vaccination and prior diagnosed infection, suggests that the observed continued ecologic decoupling of COVID-19 cases and severe outcomes is at least partly due to growing protection against severe disease from both prior infection and vaccination. With the progression of the SARS-CoV-2 pandemic globally, it is increasingly difficult to determine the clinical severity of any variant in a completely naïve individual. However, for health service planning this is less relevant than the real-world effect in populations with varying degrees of immune protection (Mefsin et al., 2022). For example, although we showed similar risk of severe hospitalization or death in the BA.4/BA.5 and BA.1 waves when adjusted for vaccination and prior diagnosed infection, the actual burden of admissions and deaths was much lower in the BA.4/BA.5 waves, with the peak 7-day moving average of admissions and deaths being 222 and 36 in the BA.1 wave vs. 66 and 9 in the BA.4/BA.5 wave. The ability to use routine data to rapidly assess the relative severity of waves caused by different lineages and variants adjusted for comorbidities, vaccination and prior infection has been especially valuable for local health service planning (Davies et al., 2022).

To our knowledge, this is one of the first comparisons of clinical severity of BA.4/BA.5 infections with previous variants with relatively complete adjustment for comorbidities and vaccination among all diagnosed cases. Nonetheless, this type of data and analysis have several limitations which have been described in detail previously (Davies et al., 2022). These include using the time of infection as a proxy for the variant causing infection rather than actual genomic sequencing or PCR test proxies (Wolter et al., 2022) which would be more accurate and overcome challenges with comparing disease severity across waves due to differences in testing practices, treatment availability and health service pressures. Notably, testing in the BA.4/BA.5 wave was at the lowest levels since the start of the pandemic with less testing of patients with milder disease, hence we may have over-estimated disease severity in this wave. With routine data we were unable to distinguish between severe hospitalizations and deaths where the diagnosis of COVID-19 may have been incidental or contributory rather than causal, and had incomplete ascertainment of key covariates especially prior diagnosed infection due to substantial missed diagnoses, vaccinations received outside of the province or without submitting a South African identity number and undiagnosed comorbidities as we can only adjust for those algorithmically identified in the WCPHDC.

In conclusion, we found similar disease severity amongst diagnosed COVID-19 cases in the BA.4/BA.5 and BA.1 periods in the context of growing immunity against SARS-CoV-2 with strong protection against severe outcomes conferred by prior infection and vaccination, especially if boosted. Ensuring that individuals at high risk of severe COVID-19 outcomes have at least three vaccine doses remains a key strategy for limiting the public health impact of further COVID-19 waves.

## Data Availability

The underlying data are de-identified pseudo-anonymised routine patient records for which patients have not consented to their de-identified data being part of publicly accessible repositories with inherent risks of re-identification. The Western Cape Department of Health and Wellness evaluates research proposals for all research in the public health sector in the Province, including those which draw on similar datasets to the current study, based on routine data, subject to standard research ethics, government approval and data governance prescripts.

## Acknowledgements

We would like to acknowledge all patients in the Western Cape and to thank the Western Cape Department of Health Provincial Health Data Centre (WCPHDC), the South African National Department of Health and the Electronic Vaccine Data System, the Western Cape Department of Health COVID-19 Outbreak Response Team, the Western Cape Communicable Disease Control sub-directorate and Western Cape health care workers involved in the COVID-19 response for their contributions to this report.

## References

Bingham J, Cable R, Coleman C, Glatt TN, Grebe E, Mhlanga L, et al. Estimates of prevalence of anti-SARS-CoV-2 antibodies among blood donors in South Africa in March 2022. Res Sq 2022. doi: 10.21203/rs.3.rs-1687679/v1.

Boulle A, Heekes A, Tiffin N, Smith M, Mutemaringa T, Zinyakatira N, et al. Data Centre Profile: The Provincial Health Data Centre of the Western Cape Province, South Africa. International Journal of Population Data Science 2019;4(2).

Callaway E. What Omicron’s BA.4 and BA.5 variants mean for the pandemic. Nature 2022;23 June 2022. doi: https://doi.org/10.1038/d41586-022-01730-y.

Davies MA, Kassanjee R, Rousseau P, Morden E, Johnson L, Solomon W, et al. Outcomes of laboratory-confirmed SARS-CoV-2 infection in the Omicron-driven fourth wave compared with previous waves in the Western Cape Province, South Africa. Trop Med Int Health 2022; 27(6):564–73.

Hussey H, Davies MA, Heekes A, Williamson C, Valley-Omar Z, Hardie D, et al. Assessing the clinical severity of the Omicron variant in the Western Cape Province, South Africa, using the diagnostic PCR proxy marker of RdRp target delay to distinguish between Omicron and Delta infections - a survival analysis. Int J Infect Dis 2022;118:150–4.

Iketani S, Liu L, Guo Y, Liu L, Chan JF, Huang Y, et al. Antibody evasion properties of SARS-CoV-2 Omicron sublineages. Nature 2022;604(7906):553–6.

Khan K, Karim F, Ganga Y, Bernstein M, Jule Z, Reedoy K, et al. Omicron sub-lineages BA.4/BA.5 escape BA.1 infection elicited neutralizing immunity. medRxiv 2022. doi: 10.1101/2022.04.29.22274477.

Kimura I, Yamasoba D, Tamura T, Nao N, Oda Y, Mitoma S, et al. Virological characteristics of the novel SARS-CoV-2 Omicron variants including BA.2.12.1, BA.4 and BA.5. bioRxiv 2022. doi: 10.1101/2022.05.26.493539.

Lewnard JA, Hong VX, Patel MM, Kahn R, Lipsitch M, Tartof SY. Clinical outcomes associated with SARS-CoV-2 Omicron (B.1.1.529) variant and BA.1/BA.1.1 or BA.2 subvariant infection in southern California. Nat Med 2022. doi: 10.1038/s41591-022-01887

Madhi SA, Kwatra G, Myers JE, Jassat W, Dhar N, Mukendi CK, et al. Population Immunity and Covid-19 Severity with Omicron Variant in South Africa. N Engl J Med 2022; 386(14): 1314–26.

Mefsin Y, Chen D, Bond HS, Lin Y, Cheung JK, Wong JY, et al. Epidemiology of infections with SARS-CoV-2 Omicron BA.2 variant in Hong Kong, January-March 2022. medRxiv doi: 10.1101/2022.04.07.22273595

Network for Genomic Surveillance in South Africa. SARS-CoV-2 Sequencing Update 10 June 2022; 2022. Available from: https://www.nicd.ac.za/wp-content/uploads/2022/06/Update-of-SA-sequencing-data-from-GISAID-10-June-2022.pdf. [Accessed 26 June 2022].

Sun K, Tempia S, Kleynhans J, von Gottberg A, McMorrow ML, Wolter N, et al. SARS-CoV-2 transmission, persistence of immunity, and estimates of Omicron’s impact in South African population cohorts. Sci Transl Med 2022:eabo7081. doi: 10.1126/scitranslmed.abo7081.

Tegally H, Moir M, Everatt J, Giovanetti M, Scheepers C, Wilkinson E, et al. Continued Emergence and Evolution of Omicron in South Africa: New BA.4 and BA.5 lineages. medRxiv 2022. doi: 10.1101/2022.05.01.22274406.

UK Health Security Agency. SARS-CoV-2 variants of concern and variants under investigation in England Technical briefing 43; 2022. Available from: https://assets.publishing.service.gov.uk/government/uploads/system/uploads/attachment_data/file/1085404/Technical-Briefing-43.pdf. [Accessed 26 June 2022.

Viana R, Moyo S, Amoako DG, Tegally H, Scheepers C, Althaus CL, et al. Rapid epidemic expansion of the SARS-CoV-2 Omicron variant in southern Africa. Nature 2022; 603(7902): 679–86.

Western Cape Department of Health. Western Cape Burden of Disease Rapid Review Update 2019; 2020. Available from: https://www.westerncape.gov.za/assets/departments/health/burden_of_disease_report_2020.pdf. [Accessed 1 March 2020].

Western Cape Department of Health in collaboration with the National Institute for Communicable Diseases SA. Risk Factors for Coronavirus Disease 2019 (COVID-19) Death in a Population Cohort Study from the Western Cape Province, South Africa. Clinical Infectious Diseases 2020;73(7):e2005–e15.

Wolter N, Jassat W, Walaza S, Welch R, Moultrie H, Groome M, et al. Early assessment of the clinical severity of the SARS-CoV-2 omicron variant in South Africa: a data linkage study. Lancet 2022;399(10323):437–46.

